# COVID Testing in the Workplace: Return to Work Testing in an Occupational Cohort

**DOI:** 10.1101/2022.02.09.22270653

**Authors:** Robby Sikka, Anne L. Wyllie, Prem Premsrirut, Ethan M. Berke

## Abstract

**Background:** The Omicron SARS-CoV-2 variant resulted in significant community-based transmission. Numerous occupational settings have employed surveillance testing strategies to minimize occupational exposure and return workers safely to work following isolation.

**Methods:** From an occupational COVID-19 testing program, we obtained longitudinal (February 2021-January 2022) saliva-based RT-qPCR results and starting December 27th, 2021, daily on-site molecular over-the-counter (OTC) nasal swab-based isothermal nucleic acid amplification test (molecular OTC; Cue Health COVID-19 test) results. We quantified the fraction of tests with PCR cycle threshold (Ct) values <30 on each day post detection from suspected and confirmed Omicron infections (n=37), compared results to molecular OTC testing, and measured workplace and household transmission. We evaluated return-to-work timing using a post-isolation, two-test threshold of Ct >30, or two negative molecular OTC tests over a 24 hour period, or a single PCR test >30 plus negative molecular OTC test.

**Results:** From the paired testing cohort, 37 (48%) individuals tested positive; all 37 were vaccinated. All individuals tested positive ≤1 day after a previous negative test, and 19 (51.3%) remained PCR-positive with Ct values <30 at day 5. While 3 (8.1%) remained PCR-positive with a Ct value <30 on day 10, no individuals remained PCR-positive on day 12. The average time to PCR clearance/return-to-work was 7.94 days (median=9.5 days). Time to clearance for those boosted (n=8; 7.75 days) and those not yet boosted (8.04 days) did not differ (*p*=0.49). Peak viral load measured by PCR was 1.97 days from the initial positive test. There were no cases of transmission after returning to work.

**Conclusions:** A large percentage of individuals remain contagious at day 5 post first positive test based on serial PCR testing and can continue until day 12. Early discontinuation of isolation can utilize a two test framework separated by 24 hours. Rapid onsite tests may be useful.

## Introduction

The introduction of the SARS-CoV-2 Omicron variant to the United States has led to significant outbreaks across the nation. Due to Omicron’s increased transmissibility, partial immune evasion, and evidence suggesting potentially shorter generation interval relative to previous variants, the virus dynamics of this particular variant have significant policy and public health guidance implications.^1^ Isolation periods which do not correspond to infectious periods may create scenarios where individuals return to work while at risk of continuing to shed transmissible levels of virus.^2,3^ Testing in corporate settings has the potential to reduce additional community introduced transmission. Scenarios where corporate testing can be performed in a reproducible manner, has the added benefit of potentially allowing businesses to remain open. The concept of “Test to Stay” has been advocated in schools and has utilized rapid antigen tests or rapid molecular tests, but testing to exit isolation has been less described.^4,5^ Use of these tests in the corporate setting has not been described to date, nor has its use in guiding return to work. Obtaining cycle threshold (Ct) values from PCR testing can guide return to work and management of isolation. While it remains unclear on the exact Ct value associated with transmissibility, a proxy has been used in many settings to determine relative infectivity.

We describe SARS-CoV-2 Omicron variant viral dynamics in individual infections in a corporate setting as well as return-to-work results from this cohort. Paired saliva-based PCR tests and nasal swab-based loop-mediated isothermal amplification test (molecular OTC) tests were utilized in this prospective cohort to detect workplace transmission. Additionally, vaccination status, symptomatology, and contact tracing results are described as these results can help inform optimal isolation durations, and testing strategies for other corporate settings.

## Methods

### Study design

The study cohort consisted of individuals who participated in a corporate COVID-19 testing program for a company with offices in New York City, NY, and Los Angeles, CA. All samples were collected between February 8, 2021, and January 15, 2022. From February 16, 2021-June 7, 2021, a four day in-person work week was employed with Monday PCR testing and a second PCR test was administered to asymptomatic individuals on their last day of work of the week. Testing was performed for any symptomatic individual. From June 7, 2021-December 15, 2021, a full 5 day in-person work week was employed with twice-weekly PCR (first and last day of work week) testing. From December 15 to December 27, daily PCR testing was employed, and from December 27 to January 15, daily PCR and molecular OTC tests were performed 6 days a week. From January 10 to 25, a combined oral and anterior nasal swab was used for molecular OTC testing. This protocol utilized a self-administered oral swab first to both tonsillar pillars, and use of that same swab to both nostrils.

Individuals self-collected saliva samples that were tested same-day using SalivaDirect, a saliva-based, extraction-less PCR test.^6^ Clinical anterior nares swabs were self-administered by individuals daily and tested immediately by molecular OTC tests using Cue Health (San Diego, CA). PCR Ct values were converted to virus genome equivalents using a standard curve (Supplementary Table 1). Viral proliferation duration (time from first possible detection to peak viral concentration), viral clearance duration (time from peak viral concentration to clearance of acute infection), duration of acute infection (proliferation duration plus clearance duration), and peak viral concentration for each person were all calculated from saliva PCR results.

Return-to-work testing utilized both PCR and molecular OTC testing. Beginning on day 4 post first positive test, criteria to return to work required being asymptomatic for at least 24 hours, followed by (A) two consecutive PCR tests with a Ct value >30, performed 24 hours apart, with a downward trend in viral load, or (B) two consecutive negative molecular OTC tests, performed 24 hours apart (Figure 1). Alternatively, after being asymptomatic for at least 24 hours, an individual could have a single PCR test with a Ct value >30, followed by a negative molecular OTC test 24 hours later. A time-based return to work was offered to employees, requiring a 10 day isolation period following the first positive test, with a single negative molecular OTC test to return. No employees utilized the time-based return to work protocol. Additional mitigation strategies employed included a vaccine mandate, required masking using a KN95 mask, limiting guests at the workplace, and all meals eaten outside. Testing was offered to family and household members and was utilized for contact tracing purposes.

**Figure 1.**
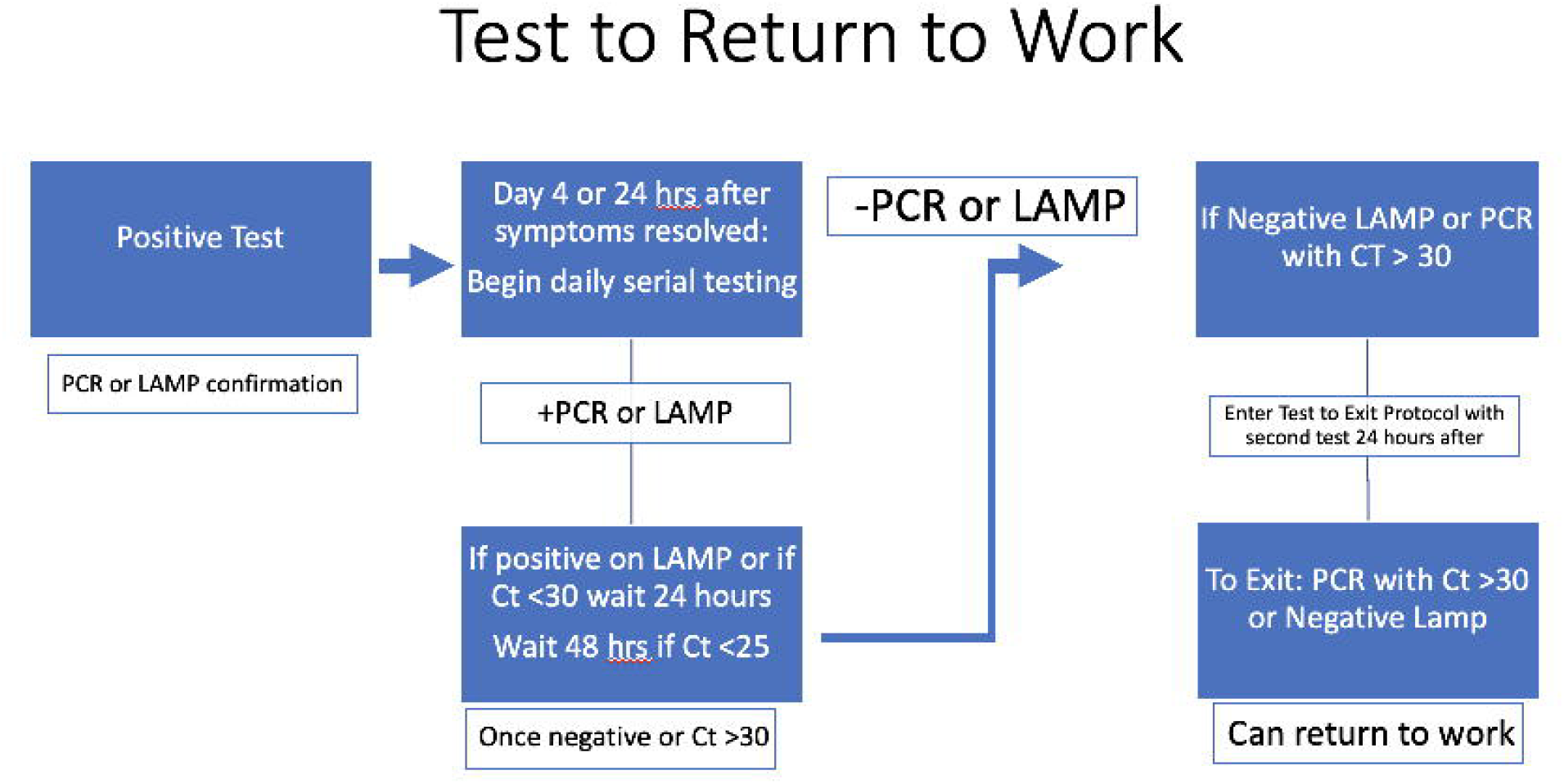
Proposed ‘return-to-work’ strategy using PCR and/or molecular OTC testing. Starting on day 4 post first positive test or at least 24 hours following resolution of symptoms, individuals can exit quarantine and return-to-work with two consecutive PCR tests with a Ct value >30, performed 24 hours apart, with a downward trend in viral load, or two consecutive negative molecular OTC tests, performed 24 hours apart, or being asymptomatic for at least 24 hours followed by a single PCR test with a Ct value >30 then a negative molecular OTC test 24 hours later.

### Study oversight

In accordance with the guidelines, this work with de-identified samples was approved for research by the SUNY Downstate Institutional Review Board & Privacy Board under the title “Pilot program for instituting massive COVID19 surveillance screening in schools and the workplace (IRB#1603504-6).”

### Variant identification

PCR-positive samples were tested with an additional RT-qPCR assay to indicate likelihood of Delta or Omicron variants by the presence or absence of S-gene detection spanning the S 69/70 deletion found in Omicron lineage BA.1 (the dominant Omicron lineage during the study period). For whole genome sequencing, RNA was extracted and confirmed as SARS-CoV-2 positive by RT-qPCR with the Thermo Fisher TaqPath SARS-CoV-2 assay. Next Generation Sequencing with the Illumina COVIDSeq ARTIC primer set2 was used for viral amplification.

### Statistical analysis

A Kaplan Meier analysis estimated the median time from testing PCR positive to Ct value >30, the cut-off of infectiousness used for the study. We defined PCR positivity as the first specimen collected with detection of SARS-CoV-2 RNA at a Ct <35. Analyses were conducted in R 4.1.2.

## Results

In total, 6645 molecular OTC and PCR tests were completed over the study period. 5994 PCR tests were conducted and 651 paired molecular OTC tests. 78% of tests were conducted as surveillance testing to minimize the number of SARS-CoV-2-positive individuals entering the facility, and reducing the risk of possible transmission, and 22% of tests were for permitting the return of employees sooner than the 10 day quarantine period that was otherwise enforced at a time. The most commonly used return-to-work approach was two negative molecular OTC tests, performed on-site (20 individuals).

From the total population of 76 adult individuals 10 tested positive between February 16, 2021-December 13 2021 (13.1%). From December 13, 2021-January 15, 2022, 37 individuals tested positive (48.8%), of which 6 (7.9%) were re-infections. All 37 individuals were vaccinated; 8 individuals were eligible, and had obtained a booster and 3 individuals were eligible and had not obtained a booster. Of the 37 individuals who tested positive in the later study period, 35 (94.6%) had mild-to-moderate symptoms and 2 (5.4%) reported symptoms persisting >3 weeks after infection. After testing positive ≤1 day after a previous negative or inconclusive test, 19 (51.3%) remained PCR-positive with Ct values <30 at day 5, 13 (35.1%) at day 6, 10 (27.0%) at day 7 and 7 (18.9%) at day 8 post-detection and 6 (16.2%) on day 9. While 3 (8.1%) remained PCR-positive with a Ct value <30 on day 10, no individuals tested PCR-positive on day 12. The average time to PCR clearance was 7.94 days, and the median time was 9.5 days (Figure 2). The average time to peak viral load as measured by PCR testing was 1.97 days from the initial positive test (Figure 3). There was no statistically significant difference in time to clearance for those boosted (7.75 days) and those not yet boosted (8.04 days) (*p*=0.49). Of the 3 individuals whose Ct values remained under 30 up until 10 days, none reported symptoms after day 6. During the initial study period prior to the first Omicron variant case, from February 16, 2021-December 15, 2021, there were a total of 208 possible work days and the work setting was fully open during 194 days (93.2% of possible days). Following Omicron, and the initiation of molecular OTC testing on December 27th, 2021, the facility has been open 100% of possible days.

**Figure 2.**
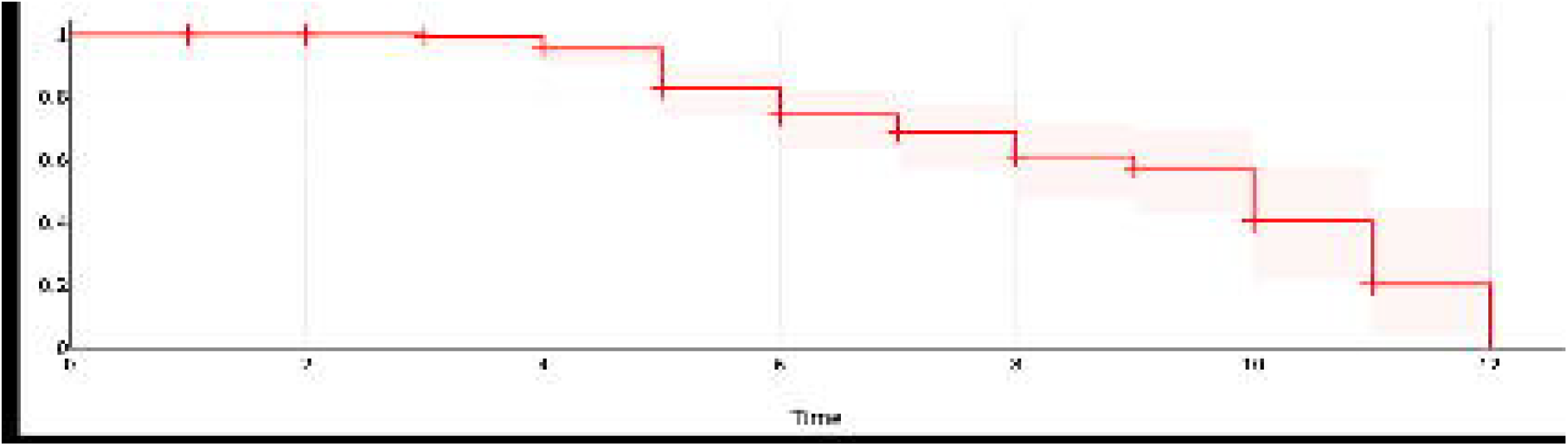
Kaplan Meier analysis estimated the median time to PCR clearance of SARS-CoV-2 was 9.5 days.

**Figure 3.**
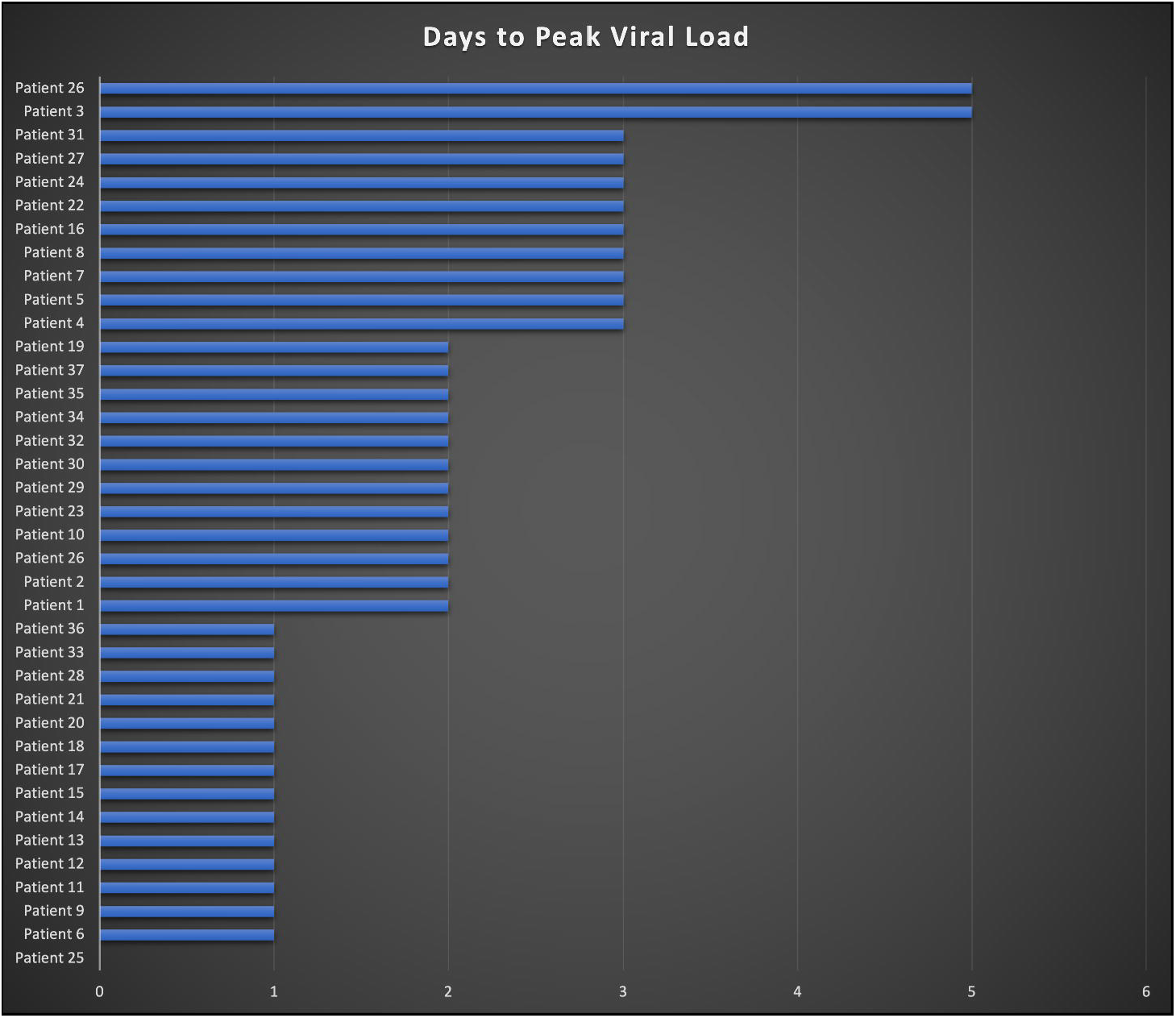
Days to peak viral load by patient. The average time to peak viral load as measured by PCR was 1.97 days from the initial positive test. Note the peak viral load for patient 25 was measured on Day 0.

For individuals who were returning to work and testing with both PCR and molecular OTC tests, individuals returned their first negative molecular OTC test approximately 2.1 days sooner than their first negative PCR test. Since the return-to-work protocol called for two consecutive negative molecular OTC tests over a 24 hour period, or a single PCR with Ct >30 plus an additional negative molecular OTC test 24 hours later, 4 individuals transitioned from molecular OTC-negative to molecular OTC-positive, which resulted in delayed return to work (Figure 5).

There was no evidence of virus transmission from any previously-positive individuals following their clearance to return to work. Prior to the first case of Omicron, there were no documented cases of workplace transmission in the study worksite. Following the emergence of Omicron, however, two cases were identified as potential workplace spread cases that may have occurred in office. Subsequent household transmission was also noted in the case of 5 individuals. Ct values from their tests within 48 hours of transmission and prior to isolation ranged from 18-30. There were no cases of transmission from any individuals who had a Ct value higher than 31 after examining transmission patterns among cases, including among household members.

In the later study period, approximately 457 matched PCR and oral-nasal swab molecular OTC tests were performed. In this setting, two individuals tested positive with oral nasal-swab molecular OTC testing one day prior to testing PCR-positive. However, two individuals tested negative by molecular OTC nasal swab testing but tested saliva-based PCR-positive with Ct values of 27 and 25. The overall concordance between saliva-based PCR and oral-nasal swab molecular OTC testing was 98.81% and the sensitivity and specificity were 92% and 99.1%, respectively. The limit of detection of the oral-nasal swab molecular OTC test equated to 36 Ct for the saliva PCR test. However, for PCR-positive saliva samples with a Ct <30, concordance with the combined oral-nasal swab molecular OTC test was 100%. Overall, oral-nasal swabs were well tolerated by the staff with no complaints, and a high rate of concordance with PCR.

## Discussion

The suitability of PCR testing in the workplace has been a point of contention throughout the pandemic. While considered the gold standard for COVID-19 diagnosis, delays in testing results during periods of high demand have resulted in the prolonged isolation of employees or extended workplace closures. Despite this, many organizations have identified the benefits of employee testing, and minimizing the chance of those positive for SARS-CoV-2 from entering facilities, particularly where the risk of COVID-19 impacting mission critical operations is high. Here, we report on a workplace testing program which demonstrates the utility of both ‘Test to Stay’ and ‘return-to-work’ testing approaches which can minimize outbreaks and maximize site operations. Daily testing during times of increased transmission and positive cases provides a ‘Test to Stay’ approach, permitting those testing negative to continue working in-person while identifying infected individuals early on, and minimizing the risk of transmission to others. Concomitantly, this daily testing program can be utilized to screen individuals as they resolve their SARS-CoV-2 infection, permitting their safe return to the workplace, avoiding unnecessarily prolonged isolation.

Following a successful workplace saliva-based PCR testing program throughout 2021, the testing protocol was adapted in response to the emergence of the SARS-CoV-2 Omicron variant and the increased number of positive cases due to its higher rate of transmissibility. In this later study period, daily PCR testing was paired with daily on-site molecular OTC testing for screening workplace employees and evaluating return-to-work strategies. A parallel design was initially adopted to ensure workplace safety and reduce the risk of transmission where unmasked activity was required, with the goal of eventually transitioning to only molecular OTC testing. Later, through comparison of PCR vs. molecular OTC results and emerging data on variable detection of Omicron by sample type,^2^ it was felt that the organization would benefit from switching to the combined oral and nasal single swab approach described in the methods. This resulted in 100% concordance with saliva PCR for all positives with Ct values <30 and for all negative tests. While oral-nasal swab-based molecular OTC testing was not compared to PCR testing and Ct values from nasal swabs alone,^7,8^ given the high concordance of oral-nasal molecular OTC to saliva PCR and little evidence of onward transmission from positive cases, the employer felt confident that this protocol allowed the early detection of pre-symptomatic cases.

On-site molecular OTC testing also enabled the organization to operate a Test to Stay protocol. Individuals were tested each day, and if negative, could continue working on-site. Over the study period more than two dozen people were considered close contacts to positive individuals at the workplace or outside of work, and despite this, only two cases of workplace transmission were noted. As a result, the workplace was able to remain open for over 90% of the 11 month study period, including when testing with only saliva-based PCR and during the Omicron surge.

Antigen tests were not used in this testing program. Early data following the emergence of Omicron raised concerns that, antigen tests could not provide the level of sensitivity, or the limit of detection, needed to ensure that a negative test meant someone was non-contagious.^2,8,9^ As such, paired molecular OTC and PCR tests were used to return employees to work. If NAAT tests are unavailable, a 3-test protocol using an antigen test may be useful to clear individuals,^10^ particularly given the likelihood of being contagious on day 10 (Ct cut-off of 30) was approximately 8% in the current study.^10,11^ Additionally, the authors felt that based on the concordance data of the combined oral/nasal swab with PCR, use of this technique would provide the most efficient testing strategy.

For returning employees to work, testing those who were SARS-CoV-2-positive starting from day 4 post their initial positive test proved beneficial, with individuals (up to 50%) starting to test negative by molecular OTC or with PCR Ct values >30 by day 5, allowing them to return on day 6 at the soonest. Notably, the testing program described utilized the routine use of PCR Ct values for analysis of positives. While labs across the U.S. do not routinely report Ct values, in this setting Ct values were available within 12 hours from sample collection. This allowed for rapid follow-up testing, and faster potential clearance in the case of false positive results from individuals with prolonged viral shedding.

Over the course of this testing program, the majority of tests (78%) were used for surveillance testing, permitting the safe entrance of employees into the workplace and making continued site operations possible. In the later study period, a large focus of testing transitioned to providing clearance for individuals who had been infected with SARS-CoV-2 to returning workers to the workplace sooner than the 10 day quarantine period that was otherwise being enforced. This two-test re-entry protocol allowed employees to instead return to work around 8 days after their initial positive test. Thus, 22% of the total testing budget enabled the average worker to return two days sooner, which depending on the workforce setting may achieve a positive return on investment. Additional work is warranted to study this economic benefit for a workplace.

This testing program provides an example of how a dynamic response to the changing course of the pandemic allowed a workplace to stay open, first through the early detection of cases, then, in response to an unexpected outbreak following the emergence of a new variant, an updated response permitted the safe return-to-work of employees. These observations demonstrate how a flexible response can keep workplaces open safely. While current recommendations permit return-to-work after 5 days post first positive test, findings from this testing program demonstrate that following infection of individuals, even in those who are vaccinated, a large proportion remain infectious well after 5 days. If not properly managed, individuals returning to work while infectious pose a risk to others in the workplace which could result in a decrease in operations or even facility closure. Rather, with serial testing after day 4, and employing a high performance on-site testing platform, employers can minimize the risk of infectious individuals returning, thereby ensuring continued worksite operations for optimal productivity.

## Data Availability

All data produced in the present study are available upon reasonable request to the authors

## Acknowledgements

We thank the COVID-19 Sports and Society Working Group and Occupational COVID Testing Managers and Staff who coordinated daily testing,

## Supplementary Material

Standardized viral concentration curve used for PCR testing

