## Supplementary figures and images for "COVID Testing in the Workplace: Return to Work Testing in an Occupational Cohort"

### Supplemental Table

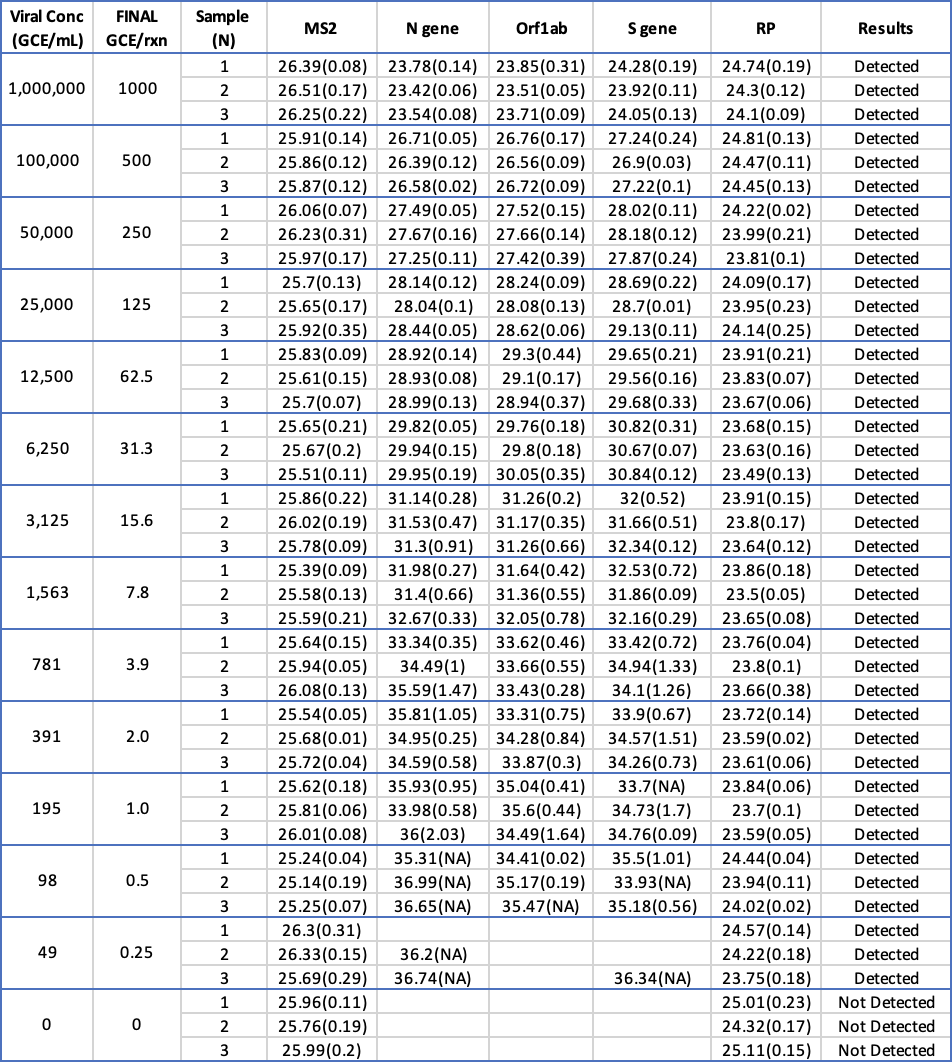
